# Integrated analysis of cell-free DNA for the early detection of cancer in people with Li-Fraumeni Syndrome

**DOI:** 10.1101/2022.10.07.22280848

**Authors:** Derek Wong, Ping Luo, Leslie Oldfield, Haifan Gong, Ledia Brunga, Ron Rabinowicz, Vallijah Subasri, Clarissa Chan, Tiana Downs, Kirsten M Farncombe, Beatrice Luu, Maia Norman, Jenna Eagles, Stephenie Pederson, Johanna Wellum, Arnavaz Danesh, Stephenie Prokopec, Eric Zhao, Nadia Znassi, Bernard Lam, Kayla Marsh, Yogi Sundaravadanam, Dax Torti, David Malkin, Raymond H Kim, Trevor J Pugh

**Affiliations:** Princess Margaret Cancer Centre, University Health Network, Toronto, Ontario, Canada; The Hospital for Sick Children, University of Toronto, Toronto, Ontario, Canada; Department of Medical Biophysics, University of Toronto, Toronto, Ontario, Canada; Vector Institute, Toronto, Ontario, Canada; Toronto General Hospital Research Institute, Toronto, Ontario, Canada; Ontario Institute for Cancer Research, Toronto, Ontario, Canada

## Abstract

Despite advances in cancer therapeutics, early detection is often the best prognostic indicator for survival (*1*). People with Li-Fraumeni syndrome harbor a germline pathogenic variant in the tumor suppressor gene *TP53* (*2*) and face a near 100% lifetime risk of developing a wide spectrum of, often multiple, cancers (*3*). *TP53* mutation carriers routinely undergo intensive surveillance protocols which, although associated with significantly improved survival, are burdensome to both the patient and the health care system (*4*). Liquid biopsy, the analysis of cell-free DNA fragments in bodily fluids, has become an attractive tool for a range of clinical applications, including early cancer detection, because of its ability to provide real-time holistic insight into the cellular milieu (*5*). Here, we assess the efficacy of a multi-modal liquid biopsy assay that integrates a targeted gene panel, shallow whole genome, and cell-free methylated DNA immunoprecipitation sequencing for the early detection of cancer in a cohort of Li-Fraumeni syndrome patients: 196 blood samples from 89 patients, of which 26 were pediatric and 63 were adults. Our integrated analysis was able to detect a cancer-associated signal in 79.4% of samples from patients with active cancer, a 37.5% – 58.8% improvement over each individual analysis. Through analysis of patient plasma at cancer negative timepoints, we were able to detect cancer-associated signals up to 16 months prior to occurrence of cancer as detected by conventional clinical modalities in 17.6% of *TP53* mutation carriers. This study provides a framework for the integration of liquid biopsy into current surveillance methods for patients with Li-Fraumeni syndrome.

## Introduction

Li-Fraumeni syndrome (LFS; OMIM 151623) is a highly penetrant hereditary cancer disorder caused by a pathogenic germline variant in the tumor suppressor gene *TP53* (*2*)(*6*). LFS patients have an estimated life-time risk of ∼75% in males and ∼100% in females of developing at least one cancer (*3, 7*), the most common being soft tissue sarcoma, osteosarcoma, brain tumors, breast cancer, and adrenocortical carcinoma (*8, 9*). However, the spectrum of at-risk cancers for LFS patients is much wider (*10*). Approximately 50% of LFS patients will develop multiple cancers (*3*) with studies showing that a younger age of diagnosis for the first cancer (*11*) and previous treatment with radiation therapy (*12*) as factors contributing to the development of subsequent cancers.

Due to their high risk of developing cancer, intensive surveillance with frequent diagnostic imaging, physical examination, and blood tests are recommended for all LFS patients; these have been shown to detect cancers earlier and improve patient outcomes (*4, 13*). Despite this, ∼20% of individuals with LFS are non-compliant, citing several negatively contributing psychosocial factors such as inconvenience, cost, logistical barriers, anxiety, and physical and emotional exhaustion (*14, 15*). Cell-free DNA (cfDNA) analysis, or “liquid biopsy”, is an emerging technology that may help to alleviate logistical barriers and complement or even replace current screening modalities. Liquid biopsy relies on the identification of tumor-associated genetic alterations or signatures (circulating tumor DNA, [ctDNA]) using DNA fragments released into the blood plasma and is an attractive biomarker due to its non-invasive collection, sensitivity, and breadth of analysis types (*5*).

With the development of cfDNA sequencing and analysis techniques, a combination of ultra-deep targeted panel sequencing (TS, ∼20,000X) and error suppression methods have detected variant allele fractions as low as 0.1% across a variety of cancer types (*16, 17*). Tumor-associated chromosomal aberrations can also be reliably detected at a lower limit of 3% of the circulating DNA using shallow whole genome sequencing (sWGS; ∼0.1-1X) (*18*). However, both approaches rely on the presence of readily detectable genetic aberrations. More recent studies have shown that ctDNA fragmentation is often shorter compared to healthy cell cfDNA fragmentation (*19*) and can provide complementary information agnostic of genetic alterations (*20*). Another field of advancement in liquid biopsy is the detection of cancer-associated methylation signatures using cell-free methylated DNA immunoprecipitation sequencing (cfMeDIP-seq) which enriches for 5-methylcytosine (*21*). DNA methylation at CpG sites is an essential component of cell identity in both normal and cancer cells and is conserved when released into circulation. Thus, enrichment of methylation signatures can be used to profile cancer-associated signatures (*22, 23*). The wide scope of analyses, non-invasive collection, and lack of need for specialty medical instruments make liquid biopsy an attractive tool for the monitoring of patients with high-risk hereditary cancer syndromes (HCS) such as LFS.

In this study, we present a cohort of 196 blood samples collected from 89 LFS patients. Using TS, sWGS, and cfMeDIP-seq, we profiled the cfDNA landscape and developed a customized multi-modal approach using genome, fragmentome, and methylome analyses to detect cancer-associated signal across a wide spectrum of cancers.

## Results

### Patient Cohort

A total of 196 blood samples were collected from 89 patients (pediatric = 26, adult = 63) recruited from clinics at the Princess Margaret Cancer Centre (n = 57) and the Hospital for Sick Children (n = 32) of which 53 of these patients (102 samples) belonged to 21 LFS families (Supplemental Figure 1A). Patients were between the ages of 1 – 67 years at the time of blood collection (Figure 1A). Male patients (n = 25; samples = 50) were found to be disproportionately less represented within the adult population compared to females (n = 64; samples = 146). Blood samples were collected from LFS patients with no known cancer at the time of blood draw (cancer negative) that included patients who have never had cancer (“LFS Healthy”) and patients with a history of cancer (“LFS Past Cancer”), and from LFS patients with active cancer (cancer positive; Figure 1B). All patients were confirmed to have been diagnosed with LFS through clinical germline testing of *TP53* (Figure 1C). A median of three serial samples (range = 2 – 11) were collected from 44 LFS patients (14 pediatric, 30 adult) of which 18 transitioned from cancer negative to cancer positive (forward; n = 8) or cancer positive to cancer negative (reverse; n = 10), termed phenoconverters (Figure 1D). The remaining patients only had one sample timepoint (Supplemental Figure 1B). We collected 38 samples from 27 LFS patients with known active cancer at the time of blood collection which included breast (n = 9), soft tissue sarcoma (n = 4), osteosarcoma (n = 3), glioma (n = 2), bladder (n = 2), prostate (n = 2), adenocarcinoma (appendiceal, n = 1), adrenocortical carcinoma (n = 1), chondrosarcoma (n = 1), endometrial (n = 1), leukemia (n = 1), lung (n = 1), and lymphoma (n = 1; Figure 1E). Within cancer-negative patients (n = 62), 22 had a previous history of cancer (LFS Past Cancer), and 40 did not (LFS Healthy). In addition to LFS patients, plasma samples were also collected from 29 healthy non-carrier control patients (“Healthy Controls”).

**Figure 1:**
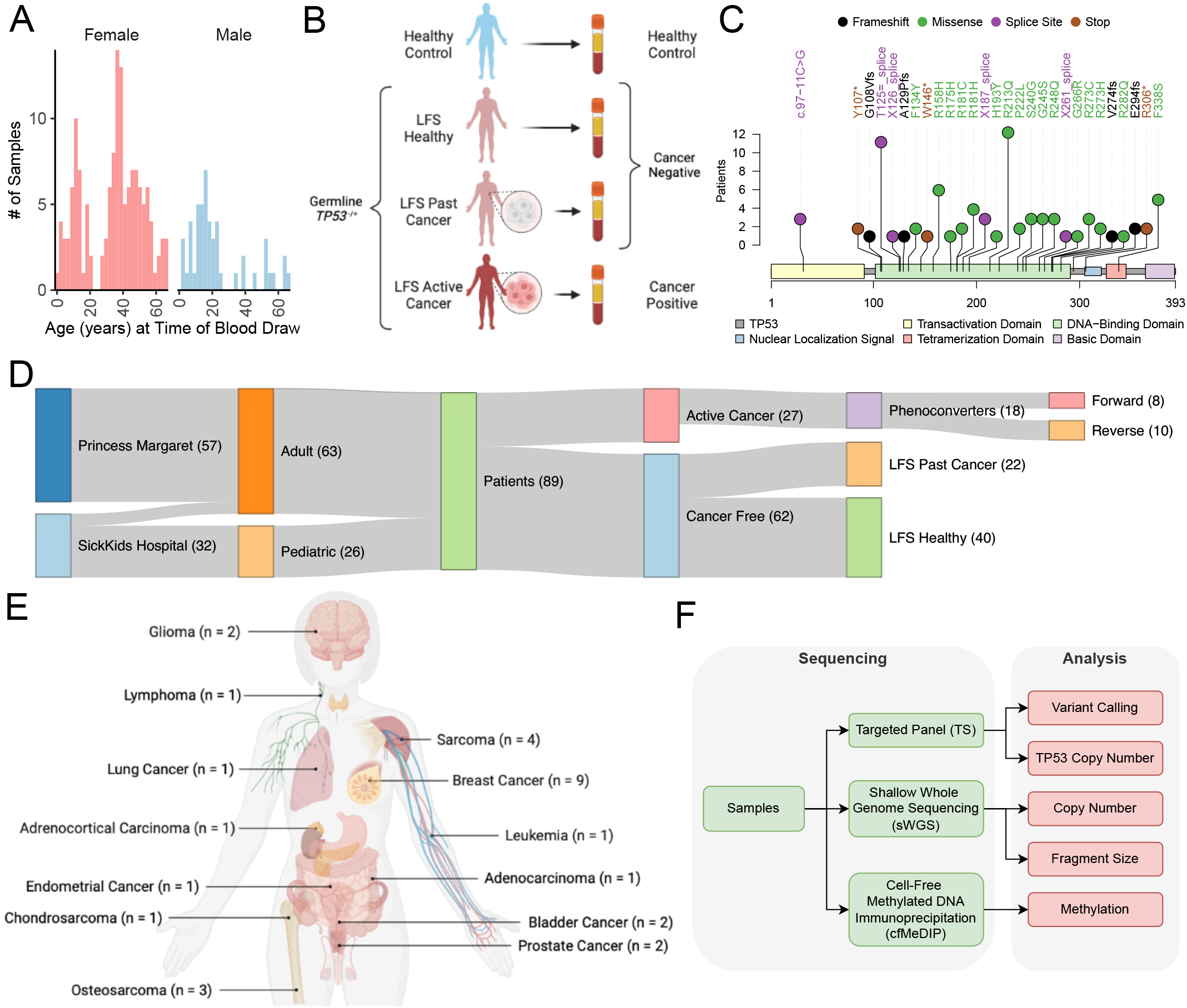
A) Histogram of LFS patient age separated by sex. B) Lollipop diagram of germline *TP53* mutations represented in our cohort C) Sankey diagram of patients in our cohort D) Location and number of cancer types clinically diagnosed in our LFS patient cohort E) Diagram showing sequencing and analysis workflow Figures 1A and 1C were Created with BioRender.com

DNA extraction yielded a median of 7.94 ng of DNA/mL of plasma (sd = 6.16) with no differences detected between pediatric (median = 8.47, sd = 5.68) and adult patients (median = 6.67, sd = 6.49) or between male (median = 8.37, sd = 4.50) and female (median = 7.23, sd = 6.68) sexes (Supplemental Figure 1C). However, due to the lower volume of blood collected from pediatric patients, total DNA yields were much lower in pediatric patients (median = 14.07 ng, sd = 10.84) compared to adult patients (median = 67.60 ng, sd = 69.98; Supplemental Figure 1D and 1E). In total, 23 samples (pediatric = 10, adult = 13) did not yield > 10ng of DNA and did not proceed with sequencing. The remaining 173 samples were submitted for a combined TS, sWGS, and cfMeDIP sequencing protocol (Figure 1E and Supplemental Figure 1F).

### Detection of somatic *TP53* mutations and copy number alterations in cell-free DNA

To determine if somatic mutations could be detected in the cfDNA of LFS patients, we performed targeted panel sequencing (TS) on 105 plasma samples from 67 patients (19 patients with serial samples). Of these samples, 26 were cancer positive, and 77 were cancer negative. A total of 70 plasma samples did not yield > 40 ng of DNA and were excluded from TS (48 pediatric, 22 adult) and 2 samples failed sequencing. Using germline variant identification, we identified 94 germline *TP53* variants (SNV/indel). In samples where an SNV/indel germline variant was not detected (n = 9), we performed targeted panel copy number analysis using PanelCNmops (*24*) and VisCap (*25*) which identified germline *TP53* copy number variants in 7 patients (Supplemental Figure 2A). Comparison between the two panel-based copy number methods resulted in high concordance (R^2^ = 0.58; Supplemental Figure 2B). In one active cancer patient (LFS90) with a germline *TP53* exon 2-9 duplication, we were not able to detect the duplication using our targeted panel copy number analysis; this may be due to contribution of a somatic *TP53* deletion event given that a shallow deletion event was detected in exons 1 and 11. In total, we identified germline *TP53* variants in 101/103 samples which were concordant with their clinical germline testing.

Amongst the 26 cancer positive samples from 20 LFS patients with active cancer, 17 were from late-stage cancers (Stage III/IV), six from early-stage (Stage 0/I/II) cancers, and three from tumors with no staging or unknown stage. TS identified 15 somatic *TP53* alterations in 11 samples from 9 patients: 8 samples from late-stage and 2 samples from early-stage cancer patients, resulting in a detection rate of 47.1% and 33.3%, respectively. Additionally, one *BRCA2*, two *PALB2*, one *MSH6*, two *PMS2*, and three *APC* somatic pathogenic variants were identified. In cancer negative samples (n = 77) from 56 cancer-free LFS patients, we identified 14 somatic *TP53* variants in 10 samples from six patients and an additional three *BRCA1*, five *BRCA2*, two *PALB2*, three *MLH1*, three *MSH2*, four *MSH6*, one *PMS2*, and three *APC* somatic pathogenic variants (Figure 2A). To further validate somatic variants and to rule out other mutagenic processes such as clonal hematopoiesis or sequencing errors, we compared the size distribution of cfDNA fragments with variants to their wildtype counterparts (*26*). Fragments with germline *TP53* variants were found to have similar size distributions compared to wildtype fragments (p = 0.187) while fragments with somatic *TP53* variants were found to have shorter fragments (p = 0.005; Figure 2B). This observation is consistent with previously published findings in sporadic cancers and provides further confidence that the somatic variants identified in our analysis are likely cancer-associated (*19*).

**Figure 2:**
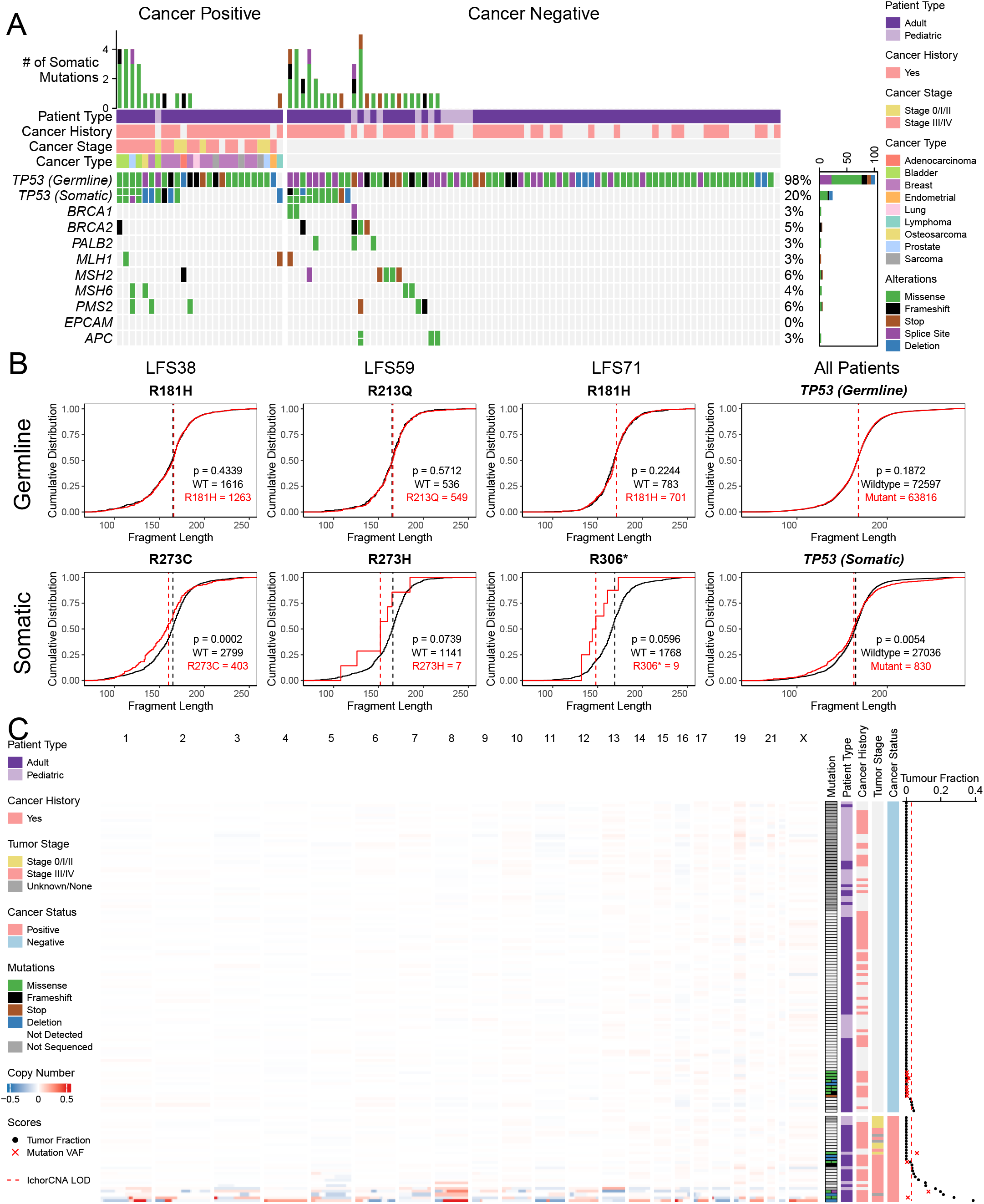
A) Oncoplot showing variants identified in our patient cohort. Germline and somatic *TP53* mutations are seperated. B) Cumulative frequency distribution graphs showing the fragment length distributions for germline and somatic *TP53* mutations identified in 3 patients. Farthest right – Cumulative frequency distribution graph of all germline and all somatic mutations identified in our cohort. Wildtype fragments are in black and germline/somatic variant fragments are in red in all graphs. P-values were calculated using the Kolmogorov–Smirnov test. C) Heatmap showing copy number alterations identified using ichorCNA.

Copy number analysis of sWGS using ichorCNA (*18*) in 134 samples (46 pediatric, 88 adult) from 85 patients (24 pediatric, 61 adult) identified 13 samples with positive tumor fractions (TF; TF > 0.03), 3/107 cancer negative samples and 10/27 cancer positive samples. Using short fragments only (90-150 bp), which has been shown to enrich for ctDNA (*27*), ichorCNA identified an additional four TF positive samples (2 cancer negative, 2 cancer positive). Comparing the two ichorCNA methodologies, we observed a high correlation between the predicted tumor fractions from full and short fragment ichorCNA analyses (R^2^ = 0.91; Supplemental Figure 2C). Unlike TS, sWGS ichorCNA analysis was not able to detect *TP53* copy number alterations (CNAs); likely due to the large (1Mb) bin size used for analysis (Supplemental Figure 2B). Robust detection of copy number alterations could be visualized using ichorCNA outputs, especially for samples from late-stage tumors with high TF (Figure 2C).

In total, 91 samples had both TS and sWGS data available − 68 cancer negative, 23 cancer positive. Within the cancer positive samples, combined TS and sWGS profiling was able to identify alterations in 14 samples (4 TS only, 5 sWGS only, and 5 both), a detection rate of 51.9%. The lack of detection in the remaining samples may be due to the relatively high TF required for robust copy number analysis (3-5% TF), lack of adequate library diversity for TS, or low shedding of ctDNA.

### LFS patients exhibit shorter cell-free DNA fragments

To further explore cancer-associated signals in the cfDNA of our cohort, we investigated the cell-free fragmentation using sWGS. Previous studies have shown an enrichment of shorter DNA fragments in patients with cancer suggesting it may be useful for detecting cancer agnostic of genomic alterations (*20, 27, 28*). We calculated the proportion of short DNA fragments (< 150 bp) in each sample and found that LFS patients, regardless of cancer status, had an increased proportion of short fragments compared to healthy controls (median = 0.176, sd = 0.027; Figure 3A) which was consistent in samples from LFS Healthy patients (n = 50, median = 0.190, sd = 0.030, p-value = 0.017; Figure 3B). Cancer positive LFS samples (n = 27, median = 0.200, sd = 0.046) additionally, had increased proportions of short fragments compared to cancer-negative LFS samples (n = 107, median = 0.190, sd = 0.035, p-value = 0.036) suggesting that the mechanism of altered fragmentation is conserved between LFS-associated cancers and sporadic cancers. This increase in short fragments observed in LFS patients was not attributable to a prior history of cancer (p-value = 0.931; Figure 3C), or age (R^2^ = 0.000; Figure 3D). We also did not observe strong correlation between the proportion of short fragments and ichorCNA tumor fraction (R^2^ = 0.098) supporting the notion that fragment size provides complimentary information agnostic of genomic alterations (Supplemental Figure 1A).

**Figure 3:**
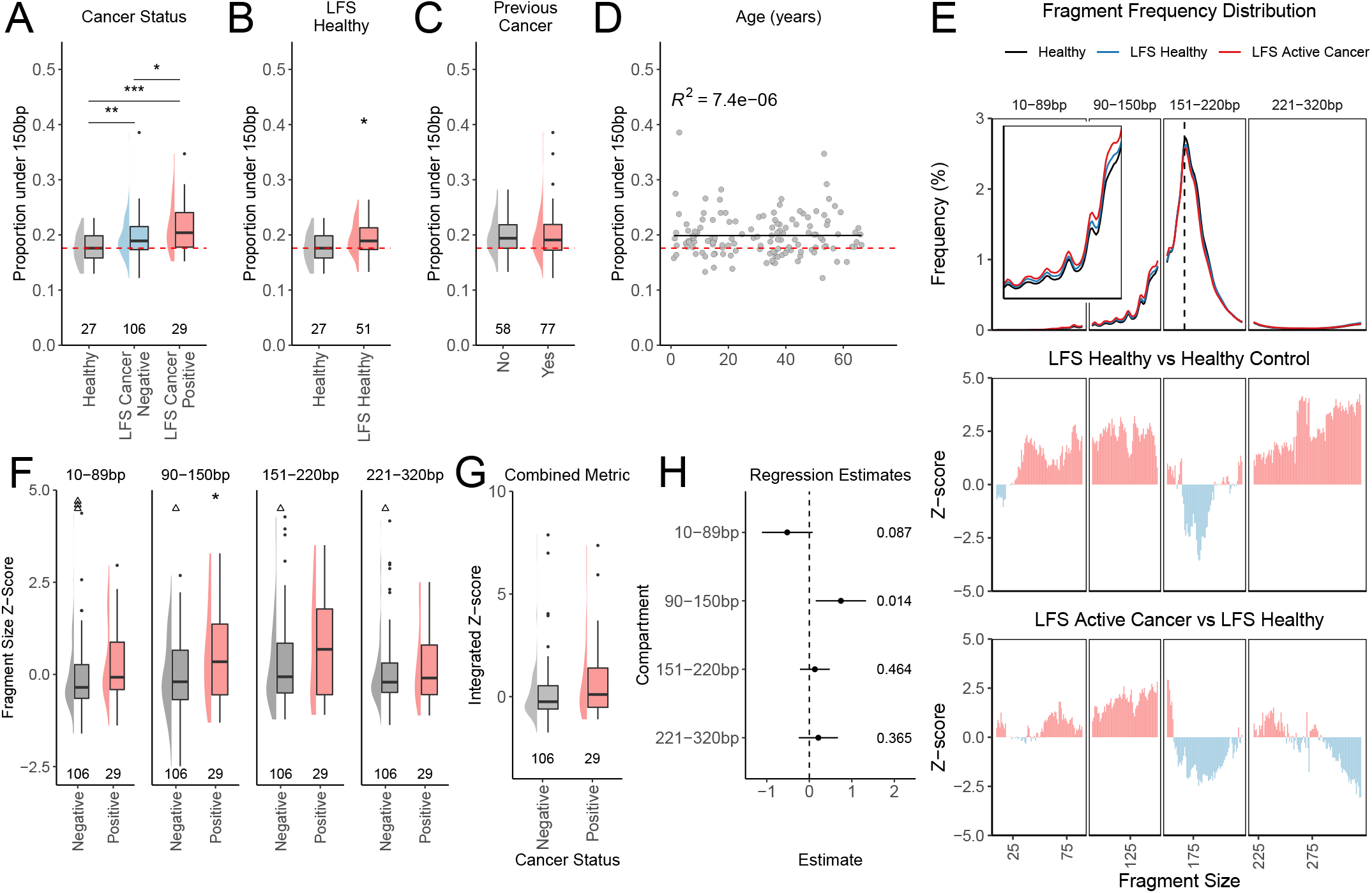
Tukey boxplots showing the distribution of the proportion of short (>150bp) fragments in healthy controls and LFS patients separated by cancer status (A), in LFS-Healthy patients (B), and cancer history (C). D) Scatter plot of the proportion of fragments >150bp and patient age at the time of blood collection. R^2^ values calculated using Pearson correlation. E) Fragment frequency distribution of samples from healthy controls (black), LFS Healthy (blue), and LFS Active Cancer patients (red; Upper). Z-scores across the fragment size distribution comparing LFS Healthy to healthy controls (Middle) and LFS Active Cancer to LFS Healthy (Bottom). Tukey boxplots showing z-scores calculated for the 10-89bp, 90-150bp, 151-220bp, and 221-320bp compartments (F) and from the integrated logistic regression model (G) between cancer negative and positive LFS samples. H) Weights and standard errors of each compartment in the integrated model. p-values are displayed on the right. p-values were calculated using two-sided Student’s t-test. * p-value < 0.05, ** p-value < 0.01, *** p-value < 0.001

To investigate differences in the distribution of fragment lengths in our cohort, we compared the median distribution of samples from healthy controls (n = 27), LFS Healthy (n = 50), and LFS Active Cancer (n = 28) patients across four fragment length compartments (10-89bp, 90-150bp, 151-220bp, and 221-320bp; Figure 3E and Supplemental Figure 3B). Compared to healthy controls, samples from LFS Healthy patients had an increased proportion of short fragments within the 10-89bp, 90-150bp and 221-320bp compartments and decreased proportions in the 151-220bp compartments (Figure 3E). In contrast, samples from LFS Active Cancer patients exhibited an increased proportion of fragments in the 90-150bp and front half of the 221-320bp compartments, and a decreased proportion of fragments in the 151-220bp compartments and later half of the 221-330bp compartments compared to samples from healthy LFS patients. This pattern of differential fragmentation suggests that the fragment length distribution of LFS patients with active cancer shifts towards shorter mono- and di-nucleosome associated fragments, consistent with sporadic cancers. Using z-scores calculated from the proportion of fragments in each compartment, only the 90-150bp compartment showed an enrichment between cancer negative and cancer positive samples (p-value = 0.025; Figure 3F). These differences in fragmentation suggest a conserved mechanism of DNA fragmentation in cancer cells versus healthy cells in LFS patients and provides support for its use as a cancer-detection method despite the differences in fragmentation observed between LFS patients and healthy controls.

To investigate whether an integrated approach would perform better than each individual compartment, we performed an integrated analysis using a logistic regression model which did not result in improved separation between cancer negative and cancer positive samples (p-value = 0.100; Figure 3G). Scores from the 90-150bp compartment were found to contribute the most to the model (estimate = 0.755, p-value = 0.013) compared to all other compartments (Figure 3H). Out of 28 cancer positive samples, we identified 7 using our integrated method compared to 6 (10-89bp), 7 (90-150bp), 8 (151-220bp), and 6 (221-320bp; Supplemental Figure 3C). The 95^th^ percentile of samples from LFS Healthy patients was used as the cut-off for detection in each compartment. In contrast, out of 106 cancer negative samples, we detected 6 (integrated), 8 (10-89bp), 5 (90-150bp), 13 (151-220bp), and 14 (221-320bp) patients. The lack of improvement from integration may be due to the high correlation between the compartments (R^2^ = 0.31 – 0.69; Supplemental Figure 3D) and suggests that the 90-150bp compartment performs best for cancer detection, consistent with previous studies (*19, 27*).

### Detection of cancer using cell-free methylation analysis

To investigate if methylation analysis can be used to detect cancer-associated methylation patterns in LFS patients, we performed cfMeDIP-seq on 142 LFS samples (32 cancer positive and 110 cancer negative) from 83 patients (60 adult, 23 pediatric). Fourteen healthy controls were also included as comparators. Evaluating QC metrics, 136 samples showed a high enrichment of methylated *Arabidopsis* spike-in controls (95.6% - 99.5%) and CpG enrichment efficiency (relH = 2.71 – 3.9, GoGE = 1.75 – 2.14; Supplemental Figure 4A). Fourteen samples were removed from downstream analysis due to low quality, and two were removed due to failed sequencing. Similar to our sWGS fragment analysis, LFS patients, independent of cancer status, showed a similar increased proportion of short fragments in our cfMeDIP-seq libraries (Supplemental Figure 4B) which was correlated to the proportion of short fragments in sWGS libraries (R^2^ = 0.82 – 0.84; Supplemental Figure 4C).

As LFS patients have previously shown differential methylation (*29*), we performed differential methylation analysis comparing healthy LFS patients and healthy controls and identified 92 differentially methylated regions (DMRs; hypermethylated; Supplemental Figure 4D). Using these DMRs, we were able to robustly classify LFS patients from healthy controls (mean AUC = 0.967, 95% CI 0.961 – 0.974; Supplemental Figure 4E).

LFS patients are at a high risk to develop a wide spectrum of different tumor types. However, due to the challenges associated with curating sufficiently large training datasets, the rarity of some LFS-associated tumor types, and the multiple n of 1 tumor types in our cohort, we first explored a pan-cancer approach. Using a universal cancer marker set from Vrba *et al*, we built a pan-cancer signature by mapping the hypermethylated CpGs to their corresponding chromosome coordinates which resulted in 1,245 DMRs (*30*). Applying the pan-cancer signature, we were able to detect cancer-associated methylation in 34 LFS samples, 11 cancer positive and 23 cancer negative (Figure 4A). Cancer positive samples included samples from patients with active breast cancer (n = 6), prostate cancer (n = 1), adrenocortical carcinoma (n = 1), osteosarcoma (n = 1), bladder cancer (n = 1), and sarcoma (n = 1). Out of a total of 16, 7, and 4 samples from late-stage (Stage III/IV), early-stage (Stage 0/I/II), and unknown or no stage cancers, respectively, we were able to detect 9 late-stage (52.9%) and 2 early-stage (28.6%) cancers using our pan-cancer approach. Within cancer negative samples that tested positive using our pan-cancer approach (n = 23), 14 (60.9%) were from patients with a history of cancer.

**Figure 4:**
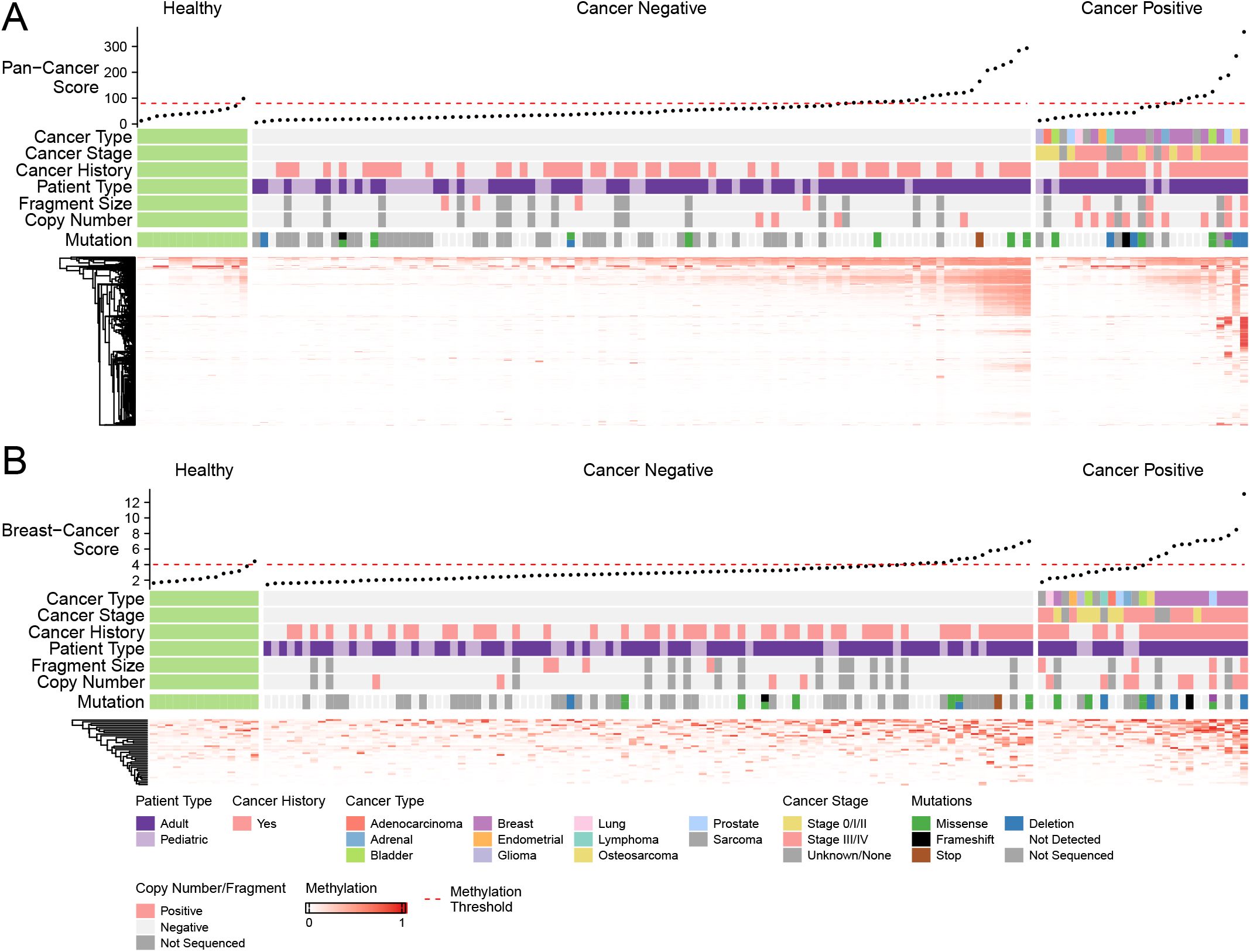
Heatmaps showing the methylation score at each methylation site in our pan-cancer (A) and breast-cancer (B) methylation signatures. A cumulative methylation score is plotted at the top. The methylation threshold was calculated using the 95^th^ percentile of the healthy control scores.

Next, as breast cancer is the most prevalent cancer type in LFS patients and in our cohort (n = 9, 44.4%), we built a breast-cancer specific methylation classifier by identifying the hypermethylated DMRs in samples from LFS patients with active breast cancer (n = 12) against samples from LFS Healthy patients (n = 48), patients with active non-breast cancer (n = 15), and healthy controls (n = 14) which resulted in a signature of 38 DMRs. A random forest classifier trained on these DMRs achieved an AUC score of 0.82 – 0.93 in predicting breast cancer samples in 10-fold cross-validation (Supplemental Figure 4F). Using this breast cancer signature, 30 LFS samples scored positive, 17 cancer negative and 13 cancer positive. Of the 13 cancer positive samples, 11 were breast cancers (8 high grade, 1 low grade, 2 unknown grade; Figure 4B). Five of the 11 breast cancer samples were not detected by the pan-cancer classifier and the one breast cancer that was not detected by either was a stage 0 ductal carcinoma in-situ that was detected on imaging due to calcification. One prostate cancer and one osteosarcoma sample also scored highly on this signature suggesting that there may be some cancer-type crossover, such as in cancers with CpG island methylator phenotype (CIMP) (*31*). Within cancer negative samples with a positive methylation score (n = 17), 12 (70.6%) samples were from survivors of a previous cancer, 7 (58.3%) of these being breast cancer. The high performance of our breast signature suggests that given robust enough training data, future studies may be able to incorporate a wide-array of cancer-type specific signatures.

### Integration of ctDNA metrics provides a holistic view of the ctDNA landscape

Individually, we demonstrated that each of our investigations were able to detect cancer associated signals, but often lacked sensitivity. To determine if an integrated approach improved our sensitivity, we compared the findings from genome, fragmentome, and methylome analyses. Out of 34 cancer positive samples, 27, 29, and 31 had corresponding TS, sWGS, and cfMeDIP-seq data, respectively, with 22 samples having all 3 analyses. Variant calling identified somatic *TP53* variants in 11 (40.7%), ichorCNA identified positive tumor fractions in 13 (38.2%), fragment size analysis identified enriched short fragments in 7 (20.6%), breast methylation identified breast-cancer associated methylation in 13 (41.9%) and pan-cancer methylation identified 11 (35.5%) cancer positive samples (Figure 5A). Combined, we were able to detect cancer-associated signal in 27 (79.4%) of all samples from patients with active cancer (Figure 5B and Supplemental figure 5A). In late-stage (III/IV) cancers, our detection rate increased to 20/21 (95.2%) while in early-stage (0/I/II) cancers, our detection rate was 4/9 (44.4%; Supplemental Figure 5B and C). Within samples from patients with active breast cancer (n = 14), methylation profiling identified 11 and was the only positive detection method in 6 samples (42.9%; Supplemental Figure 5D).

**Figure 5:**
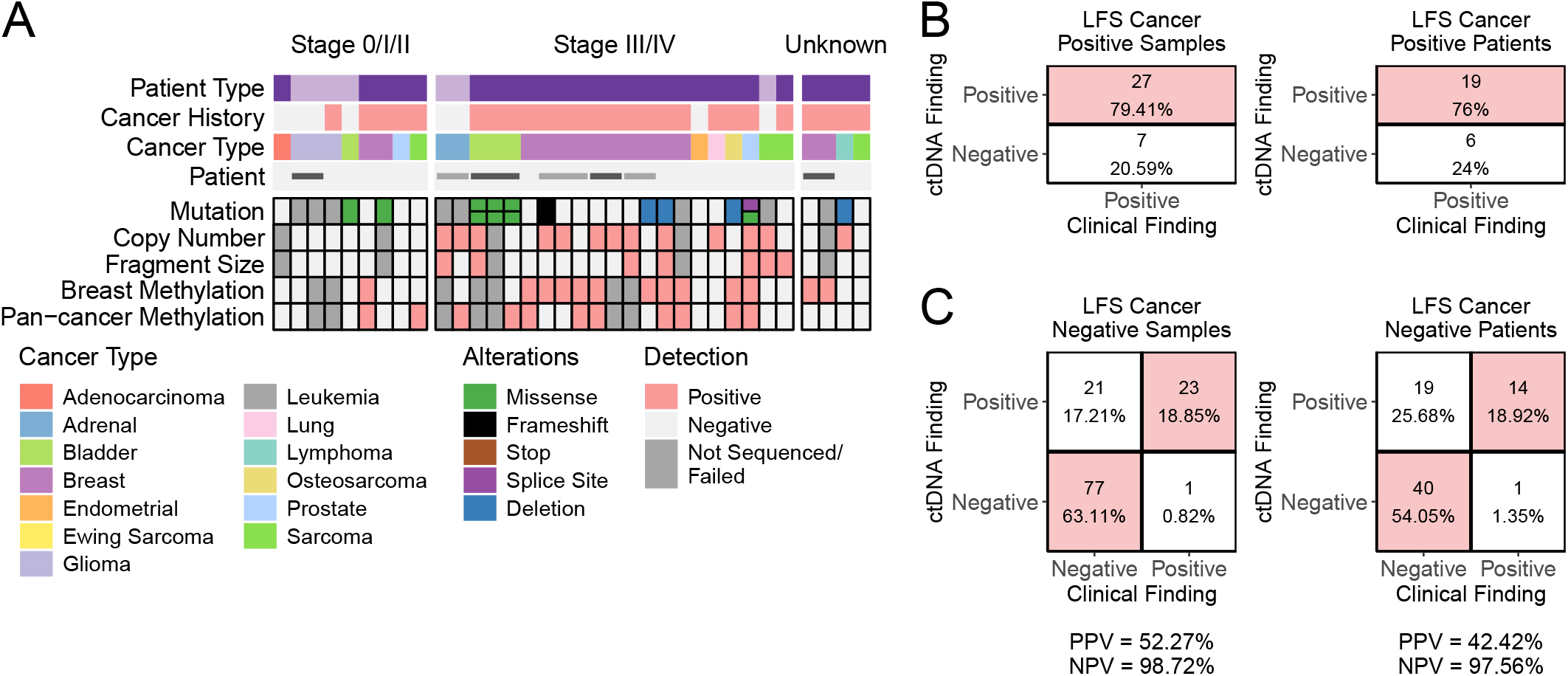
A) Comparison of cancer-associated signal detection across all analysis methods in cancer positive samples. Confusion matrixes showing detection rates for cancer positive (B) and cancer negative (C) LFS samples and patients.

In all of our analyses, several samples from cancer free patients exhibited a cancer-associated signal which may be indicative of false positives. Of 122 purported cancer-free samples, we identified 44 samples from 33 patients with any combination of somatic *TP53* variants (n = 10), cancer associated copy number alterations (n = 4) or fragmentation (n = 5), or positive breast (n = 17) or pan-cancer (n = 23) methylation scores (Supplemental Figure 5E). Cancer-associated signal was detected in only one analysis in 32/44 (72.7%) cancer free samples compared to 7/34 (20.6%) active cancer samples. Following an in-depth review of clinical data, we identified a later cancer diagnosis or suspicious finding by imaging in 14 purported cancer-free patients (23 samples; Table 1). These findings suggest that our false positive rate is 21/122 (17.2%) within samples and 25.7% (19/74) within patients. In contrast, only one patient with negative ctDNA findings was diagnosed with cancer (glioma) within 1 year following the last ctDNA timepoint analyzed. Together, within cancer negative LFS patients, our positive predictive value was 42.4% (14/33) and our negative predictive value was 97.6% (40/41; Figure 5C).

**Table 1:** Clinical findings in cancer negative patients with a molecular finding.

| Patient | Assay Positive | Months Preceding Finding | Clinical Finding |
| --- | --- | --- | --- |
| LFS5 | Breast Methylation | 4 | Osteochondroma<br>Osteosarcoma |
|  | Pan-cancer Methylation | 9 |  |
| LFS31 | Breast Methylation | 8 | Prostate Cancer |
| LFS42 | Pan-cancer Methylation | 8 | Suspicious lung nodules<br>Suspicious lymph nodes in neck |
|  | Breast Methylation | 2 |  |
|  | Breast Methylation<br>Pan-cancer Methylation | - |  |
| LFS45 | Copy Number | 11 | Lung Cancer |
|  | TP53 somatic mutation | 5 |  |
| LFS51 | Breast Methylation<br>Pan-cancer Methylation | 8 | Mucinous Adenocarcinoma and<br>Neuroendocrine Tumor of the<br>Appendix |
| LFS54 | Pan-cancer Methylation | 6 | Squamous Cell Carcinoma |
| LFS59 | TP53 somatic mutation<br>Breast Methylation<br>Pan-cancer Methylation | 15 | Lung Metastasis from<br>Leiomyosarcoma |
|  | TP53 somatic mutation<br>Breast Methylation | 8 |  |
| LFS63 | Breast Methylation<br>Pan-cancer Methylation | 6 | Liver, Thyroid, and Cervical<br>Lesions |
| LFS66 | Breast Methylation | 0 | Breast Lump |
| LFS67 | Breast Methylation<br>Pan-cancer Methylation | 10 | Basal Cell Carcinoma |
| LFS68 | TP53 somatic mutation<br>Breast Methylation<br>Pan-cancer Methylation | 21 | Schwannoma<br>Bilateral Renal AML |
|  | TP53 somatic mutation | 14 |  |
|  | TP53 somatic mutation | 3 |  |
|  | TP53 somatic mutation | 0 |  |
| LFS71 | TP53 somatic mutation<br>Breast Methylation<br>Pan-cancer Methylation | - | Tubular Adenoma (unresected)<br>Adrenal Adenoma |
| LFS73 | Pan-cancer Methylation | 10 | Metastatic Bladder Cancer |
| LFS78 | Breast Methylation<br>Pan-cancer Methylation | 12 | Liver and Bone Metastases from<br>Breast Cancer |
|  | Pan-cancer Methylation | 3 |  |

## Discussion

Here, we present a comprehensive integrated analysis of 173 liquid biopsy samples from 84 LFS patients profiled using genomic, fragmentomic, and epigenomic methods. Our approach shows and supports the power of an integrated and multi-omic ctDNA assay for the early detection of cancer in cancer predisposition syndromes such as LFS. Each assay (TS, sWGS, cfMeDIP-seq) enables analyses that measure independent biological signals which when evaluated together, can improve the overall sensitivity, specificity, and robustness of predictions.

Using TS, somatic *TP53* variants were only detected in 40.7% (11/27) of samples from patients with active cancer, suggesting that a TS-only approach may not be comprehensive enough even in patients with clinically confirmed cancer. Interestingly, we were able to show that both germline and somatic gene, and even exon-level, copy number alterations in *TP53* could be detected in the plasma. Similarly, genome-wide copy number alteration detection in the plasma, using ichorCNA which detected 38.2% (13/34) of samples from active cancer patients, was restricted by the limit of detection required for confident CNV detection (∼3-5% tumor fraction) and restricted to large events (>5Mb) (*18, 32*). While downstream bioinformatic analysis techniques have significantly increased the sensitivity of mutation detection by TS (*33, 34*), this strategy is also fundamentally limited by the reliance on idealized detection of mutations at few loci and the number of genomic equivalents available in small quantities of cfDNA (*35*). Many of our pediatric samples did not yield sufficient DNA for TS profiling, which would severely limit the use of TS in this patient population.

Using methylation profiling, we were successful in detecting 91.2% (11/12) of breast cancers using our breast cancer signature, and 40.7% (11/27) of cancer positive samples using our general cancer signature. The large discrepancy between our breast-cancer specific signature and our pan-cancer signature suggests that a targeted approach is more effective. Our pan-cancer signature was also built using methylation array data while our breast cancer specific signature was built using cfMeDIP-seq data which provides more comparable data and more granular methylation information. However, due to the availability and rarity of genome-wide methylation data, such as other cfMeDIP-seq studies or whole genome bisulphite sequencing, building cancer-type specific signatures has proven difficult especially for diagnostically challenging cancers like soft tissue sarcomas and rare tumors. Similar to a previous study, using our cfMeDIP-seq data, we were also able to identify a set of LFS-specific methylation sites which could be used to classify LFS patients from non-carrier healthy controls (AUC = 0.97, 10-fold cross-validation) (*29*). This finding may be useful for the diagnosis of LFS in patients without a detectable germline *TP53* mutation or to stratify families with LFS-like phenotypes for increased surveillance.

In this study, we found fragment size analysis to be the least sensitive but often correlated with cancer status. This lack of sensitivity may be due to the differing baseline levels of fragmentation observed between patients potentially due to genetic and lifestyle factors (*36*). Given enough longitudinal samples, patient specific fragmentomic baselines could be used to increase sensitivity rather than a general non-specific cut-off. The need for these baselines is especially useful in LFS patients due to the inherently altered cfDNA fragmentation observed in this cohort. It may also suggest that different HCS may also require patient or HCS specific baselines and controls for sensitive and robust detection.

Within cancer positive samples, combining mutation, copy number, fragment size, and methylation analyses increased our detection rate to 79.4% (27/34), a 37.5 - 58.8% increase over each individual analysis. The increase in sensitivity suggests that future clinical tests aimed towards early detection will need to integrate several analysis types that leverage different biological data to gain comprehensive insight into the cfDNA landscape. More striking, was the identification of a cancer-associated signal in 17.6% (13/74) of patients during a clinically cancer-free timepoint, wherein the participant subsequently developed a cancer or suspicious imaging finding at follow-up. In one participant’s case, our analyses were able to detect a cancer-associated signal 16 months prior to diagnosis with conventional clinical modalities. The sensitivity gained from multi-modal analyses should also be balanced with false positive rates which can lead to unnecessary and costly follow-up procedures. However, this may be tolerated in high-risk patients such as those with LFS, who already undergo routine imaging modalities that also have high rates of false positivity (*37, 38*). Healthcare providers are also enthusiastic about the potential for ctDNA in HCS but remain cognizant of false positive rates (*39*). In this study, 27% (20/74) of cancer-free patients showed a cancer-associated signal by ctDNA analysis but no cancer or suspicious imaging findings were detected. However, due to the high prevalence of cancer in this population, careful follow-up should be done as to not discount these cases as false positives. Combination of imaging surveillance and integrated ctDNA analysis may be complementary in both increasing sensitivity and decreasing the rate of false positives.

While many ctDNA studies are currently focused on singular or bimodal approaches to ctDNA detection, our study suggests that integrated multi-omic analyses should be adopted and explored further to assess the relative benefits of including additional assays, analyses, and biological information. We demonstrate that integration of multiple plasma-based analyses provides a more holistic view into the ctDNA landscape in low tumor burden settings, such as early detection. Except for cfMeDIP-seq, all analyses in this study also utilize tools that are already routinely used or can be readily and easily integrated into a clinical workflow. This study provides support for adopting multi-omic liquid biopsy for surveillance and early detection in HCS.

## Materials and Methods

### Study Design and Patient Cohort

This study was approved by the UHN institutional review board (REB# 19-6239). All patients underwent routine clinical care by board certified clinicians as per the standard-of-care. All samples were collected with informed consent for research.

### Blood Processing

Venous blood samples were collected in EDTA or Streck tubes (Streck, La Vista, Nebraska). EDTA collection was processed within 2 hours. Whole blood samples were centrifuged at 4□(1900g, 10 minutes). PBMCs were separated from plasma and stored at −80□. Isolated plasma was centrifuged a second time at 4□(16,000g, 10 minutes) to remove residual cells and debris. Purified plasma was stored at −80□until cfDNA extraction.

### DNA Extraction

DNA from blood was extracted using the QIAGEN QIAamp Circulating Nucleic Acid Kit (Qiagen, Hilden, Germany). Genomic DNA from PBMCs was extracted using the DNeasy Blood and Tissue Kit (Qiagen). After extraction, genomic DNA was sheared to similar sizes as cfDNA using an ultrasonicator (LE220, Covaris, Woburn, MA). Library preparation for TS, sWGS, and cfMeDIP-seq were performed only for samples with >40ng DNA yield. TS was excluded for samples with <40ng DNA yield. Samples with <10ng DNA yield were not processed.

### Cell-Free Methylated DNA Immunoprecipitation

cfMeDIP-seq for each sample was prepared as previously described (*21*) using 10ng of UMI ligated cfDNA library with the following deviations from the protocol. Briefly, 0.1ng of *A. thaliana* DNA methylation control package containing one methylated and one unmethylated spike-in control BAC (Diagenode, Denville, NJ) was spiked into 10ng of cfDNA library. 5% of the sample was aliquoted as a control library. The remainder underwent immunoprecipitation using monoclonal antibody targeting 5-methylcytosine (5-mc) (Diagenode, clone #33D3). Immunoprecipitated libraries and control libraries were amplified, indexed, and pooled.

### DNA Library Construction and Sequencing

Pre-capture libraries were prepared using KAPA Hyper Prep Kit (Kapa Biosystems, Wilmington, MA) and xGen Duplex Seq Adapter-Tech Access (Integrated DNA Technologies [IDT], Coralville, IA). UMI ligated cfDNA libraries were then split for each assay (TS, sWGS, cfMeDIP-seq). To generate TS libraries, hybrid capture using a custom 100kb panel of 829 probes targeting *TP53, BRCA1, BRCA2, PALB2, MLH1, MSH2, MSH6, PMS2, EPCAM*, and *APC* (Supplemental Table 1) was performed on dual indexed pre-capture libraries, then PCR-amplified. sWGS and TS libraries were indexed, pooled, and sequenced on the NextSeq 500 using 150-bp paired-end sequencing reads (2×150 bp; Illumina, San Diego, CA). sWGS libraries were sequenced to a target 1X, and TS libraries were sequenced to a target 20,000X. cfMeDIP-seq libraries were sequenced to a target 60 million clusters on the MiSeq Nano using 150-bp paired-end sequencing reads (2×150 bp; Illumina). UMI extraction and sequencing alignment to human genome reference GRCh38 was performed using Burrows-Wheelers Aligner version 0.7.12 (https://github.com/oicr-gsi/bwa) (*35*) and deduplicated using Samtools version 1.9 according to the following workflow: https://github.com/oicr-gsi/bwa. All sequencing coverage can be found in Supplemental Table 2.

### Targeted Sequencing Analysis

Aligned reads were error corrected and amalgamated using ConsensusCruncher (https://github.com/pughlab/ConsensusCruncher) (*33*) according to the following workflow: https://github.com/oicr-gsi/consensusCruncherWorkflow. Germline variant calling was performed using The Genome Analysis Toolkit version 3.8 HaplotypeCaller (Broad Institute, Cambridge, MA) (*40*). Somatic variant calling was performed using MuTect2 version 3.8 (Broad Institute) (*41*). Variant calling was performed individually on single-strand consensus sequences, duplex consensus sequences, and all unique ConsensusCruncher outputs then merged. Variant allele frequency was annotated using read support from the all_unique ConsensusCruncher output. Single nucleotide polymorphisms considered benign were removed from further analysis. Candidate pathogenic variants were manually reviewed using Integrative Genomics Viewer version 2.8.13 (Illumina). Germline variants were classified according to the 2015 American College of Medical Genetics and Genomics guidelines (*42*).

### Copy Number Variation Analysis

*TP53* copy number variants were detected using PanelCNmops (v 1.14.0) (*24*) and VisCap (https://github.com/pughlab/VisCap) (*25*). GC-correction, read count normalization, copy number prediction, and tumor burden prediction were performed using the ichorCNA tool version 0.2.0 (Broad Institute) (https://github.com/broadinstitute/ichorCNA) (*18*) using the recommended default settings. An in-house panel of normals was generated using in-house healthy blood controls (n = 31). To enrich for ctDNA, ichorCNA was performed once on all fragments and again using only short fragments (90-150bp). This was performed due to increased detection of ctDNA reported in previous studies (*27*). IchorCNA outputs were then manually reviewed. The higher predicted tumor fraction (all vs short) was used as the ichorCNA output. For all ichorCNA analyses, only predicted tumor fractions > 0.03 were considered positive (based upon the ichorCNA limit of detection).

### Fragment Size Analysis

Global fragment size distributions were calculated using Picard CollectInsertSizeMetrics (v4.0.1.2). Fragment sizes of mutations detected using targeted panels were performed using a custom script and Samtools (v1.9). Briefly, aligned reads overlapping with a mutation of interest were pulled from the bam files and then binned into wildtype and mutant reads. Reads that had alterations at the site of interest not concordant with either mutation or wildtype were discarded.

### Cell-Free Methylome Analysis

The cfMeDIP-seq FASTQ files were analyzed using an integrated pipeline known as MedRemix (https://github.com/pughlab/cfMeDIP-seq-analysis-pipeline). First, extract_barcodes.py from ConsensusCruncher (https://github.com/pughlab/ConsensusCruncher, May 19, 2021 commit) was used to extract unique molecular identifier (UMI) barcodes and remove spacers from unzipped FASTQ files (*33*). Then, alignment was performed to the human genome (genome assembly GRCh38/hg38) by BWA-MEM v0.7.17, sorted and indexed by Samtools v1.14. Next, the depth of aligned fragments generated from paired reads of cfMeDIP-seq libraries were counted within nonoverlapping 300 bp windows, along with the number of CpGs within each window. The coverage depth was modeled as a two-component mixture, with the two components representing methylated and non-methylated bins, accounting for CpG density and GC content by negative binomial regression. The mean coverage of non-methylated reads was inferred from bins with zero CpGs as a function of GC content and treated as a fixed mixture component. The mixing coefficient of methylation status and regression coefficients between the mean coverage of methylated bins and CpG density were jointly inferred using an expectation-maximization process and iterated until convergence. In this process, a posterior probability of each bin being methylated was computed.

The pancancer signatures were obtained from Vrba *et al* (*30*). The authors obtained 1,250 hypermethylated CpGs by comparing the Illumina HumanMethlation450 data of 23 types of cancer with their matched normal adjacent samples in TCGA. After mapping these CpGs to their chromosome coordinates (and then methylation bins) and removing the ones on the sex chromosomes, we obtained 1,245 DMRs in total.

To identify LFS-specific and LFS-breast cancer specific signatures, we first removed bins that overlapped with regions in the ENCODE blacklist. Bins were further filtered by removing regions frequently methylated in healthy controls samples, bins with average posterior probabilities >β in healthy control samples. Additionally, as regions that are hypermethylated in cancer samples are typically located in CpG dense regions, we removed bins with γ or more CpG sites. Using grid search, β = 0.1 and γ = 1 were chosen.

The LFS-specific signatures were identified by comparing the 52 LFS healthy samples with the 14 healthy control samples. We randomly chose 14 samples from the LFS healthy group and generated a subset by combining them with the 14 control samples. This process was repeated for 200 times, resulting in 200 balanced datasets. 100 of them were used for DMR identification, while the remaining 100 subsets were used in cross-validation. To identify DMRs, in each subset we selected the top *n*_*DMR*_ hypermethylated DMRs in the previvor group using limma R package, and only DMRs that existed in more than *k* times of the comparison were chosen. *n*_*DMR*_ and *k* were determined by grid search. Finally, *n*_*DMR*_ = 150 and *k* = 30 were chosen, and 92 DMRs were identified.

The classification performance was then evaluated on the 100 held out subsets using 10-fold cross-validation. Random forest classifiers (scikit-learn 1.0.2) were trained on each subset, and the AUROC scores were calculated.

The breast cancer DMRs were identified based on 3 groups of hypermethylated DMRs in our LFS breast cancer samples: 1) breast cancer vs LFS healthy; 2) breast cancer vs non-breast cancer; 3) breast cancer vs healthy control. The intersection of the first two groups of DMRs contains signatures specific to LFS breast cancer patients. The intersection of group 2 and 3 contains signatures specific to breast cancer or tumor samples. We then used the union of these two intersection sets as breast cancer DMRs and obtained 38 signatures. To identify DMRs in breast cancer against previvor samples, we used similar strategy as we did for identifying LFS-specific DMRs. 100 subsets were created and 185 DMRs were identified. For the other two differential methylation analyses, we did not perform subsampling since the datasets were balanced. The top *n*_*DMR*_ hypermethylated DMRs were selected. Similarly, *n*_*DMR*_ (100 for healthy control, 150 for non-breast cancer) was determined based on cross validation accuracy.

The methylation score (cancer score) of a sample is defined as the cumulation posterior probabilities of the methylation bins that correspond to the DMRs. The higher the score, the more possible that the cancer specific DMRs are methylated in the corresponding sample, which indicate the patient is more likely to be cancer positive.

### Healthy Control Cohorts

Healthy blood controls were consented and recruited with institutional approval (REB#: 19-6239) and underwent TS (n = 24), sWGS (n = 29), and cfMeDIP-seq (n = 28). All healthy control data were aligned to GRCh38 as described above and processed according to GATK best practices. Healthy control plasma analyses were performed the same as described above. Heatmap visualizations were created using the R package Complexheatmap (v2.9.4) (*43*).

## Data Availability

All data produced in the present work are contained in the manuscript

## Endnotes

## Acknowledgements

This work would not have been possible without the patients and their generous participation in this study. This work was funded by a grant from the Terry Fox Research Institute (grant number #1081) and Canadian Institutes for Health Research (DM). This study was a collaboration with the CHARM consortium (https://charmconsortium.ca) and is performed under the auspice of the LIBERATE study (NCT 03702309), which is an institutional liquid biopsy program at the University Health Network supported by the BMO Financial Group Chair in Precision Cancer Genomics (Chair held by Dr. Lillian Siu). Additional funding support for this project was made possible by the Princess Margaret Cancer Foundation and the Ontario Institute for Cancer Research (OICR). DW is supported by a Princess Margaret Cancer Center Fellowship, a Princess Margaret Cancer Digital Intelligence SPARK Award, a Canadian Institutes of Health Research Fellowship, and a Children’s Tumor Foundation Young Investigator Award. RKH is supported by The Bhalwani Family Charitable Foundation, Princess Margaret Cancer Foundation, and FDC Foundation. TJP holds the Canada Research Chair in Translational Genomics and is supported by a Senior Investigator Award from the Ontario Institute for Cancer Research and the Gattuso-Slaight Personalized Cancer Medicine Fund. DM holds the CIBC Children’s Foundation Chair in Child Health Research. We thank the staff of the OICR Genomics Program (https://genomics.oicr.on.ca) for their expertise in generating the sequencing data used in this study. OICR is supported by funding provided by the Government of Ontario. The results published here are in part based upon data generated by the TCGA Research Network: https://www.cancer.gov/tcga. We thank members of the Pugh Lab, Kim Lab, and Malkin Lab for their extensive peer editing and feedback to improve the manuscript. Lastly, we particularly thank all of the Li-Fraumeni Syndrome family members who contributed samples for the study.

## Author Contributions

DW, PL, DM, RK, and TJP designed and supervised the study. DW, PL, and TJP synthesized, interpreted, and wrote the manuscript and figures. DW performed genomic analyses. PL performed methylome analyses. AD, S Prokopec, EZ, NZ provided bioinformatics pipeline development support. JE, S Pedersen, and JW performed sample extractions and management. DM and RK recruited patients and coordinated specimen collection. LO and HG coordinated clinical data and study management. MN, RR, CC, TD, BL, KF, LB, and VS provided clinical support, clinical follow-up, and data abstraction. BL, KM, YS, DT performed, coordinated, and managed sample sequencing.

## Competing Interests

The authors declare the following financial or non-financial competing interest: TJP reports personal fees from AstraZeneca, Canadian Pension Plan Investment Board, Chrysalis Biomedical Advisors, Illumina, Merck, and PACT Pharma and grants from Roche/Genentech outside the submitted work.

## Additional Information

Supplementary Information is available for this paper.

Correspondence and requests for materials should be addressed to Trevor Pugh.

Reprints and permissions information is available at www.nature.com/reprints.

## Data and Code Availability

All sequencing data files (TS, sWGS, cfMeDIP) are deposited in the European Genome-Phenome Archive (EGA) under the accession number EGAS00001006539. Code and processed files to reproduce all analyses and figures are available at https://github.com/pughlab/LFS-early-detection-ctdna.

## Supplemental Figure Legends

Supplemental Figure 1:

A) Barplot showing the number of patients in each LFS family. The number on top denotes the number of samples from each family.

B) Sankey plot showing the breakdown of plasma samples.

C) Tukey boxplots showing cfDNA yields (ng/mL of plasma) compared between healthy controls and LFS (left), LFS pediatric and LFS adult (middle), and LFS XX and LFS XY sexes (right).

D) Tukey boxplots showing total cfDNA yield.

E) Scatter plot comparing cfDNA yield (ng/mL of plasma) with age at blood draw.

F) Upset plot showing intersect of samples with TS, sWGS, and cfMeDIP sequencing.

p-values were calculated using Student’s two-sided t-test. * p-value > 0.005, ** p-value > 0.01, *** p-value > 0.001

Supplemental Figure 2:

A) Heatmap showing amplicon based copy number predictions from PanelCNmops and VisCap normalized to cancer negative patients without germline alterations. *TP53* exons are denoted at the bottom.

B) Scatter plot comparing the copy number predictions from PanelCNmops and VisCap (Top), PanelCNmops and ichorCNA (Middle), and VisCap and ichorCNA (Bottom).

C) Scatter plot comparing the tumor fraction predictions from ichorCNA full fragment analysis and ichorCNA short fragment analysis.

R^2^ values were calculated using Pearson correlation.

Supplemental Figure 3:

A) Scatter plot showing correlation between the proportion of short (<150bp) fragments and tumor fraction predicted by ichorCNA.

B) Fragment frequency distributions comparing cancer free and active cancer LFS patients with the median healthy control (red line; Top) and cancer negative and cancer positive LFS patients with the median LFS Healthy (red line; Bottom).

C) Heatmap showing the number of standard deviations away from the LFS Healthy median for each patient (rows). Top annotation shows the frequency distribution of the LFS Healthy median. Right shows z-scores calculated for each fragment size compartment.

D) Scatter plots showing the correlation between z-scores calculated from each fragment size compartment.

R^2^ values calculated using Pearson correlation.

Supplemental Figure 4:

A) Tukey boxplots of QC metrics used to assess enrichment of methylated DNA. Dotted line represents the cut-off values for passing QC.

B) Tukey boxplots showing the proportion of fragments across three fragment size compartments in cfMeDIP-seq libraries.

C) Scatterplot showing the correlation between the proportion of short fragments in cfMeDIP-seq and sWGS libraries.

D) Volcano plot of differentially methylated regions identified in LFS patients.

AUC-ROC curve showing classification of LFS patient from non-carrier healthy controls (E) and classification of breast cancer positive LFS patients (F).

R^2^ values were calculated using Pearson correlation. p-values were calculated using Student’s two-sided t-test. * p-value > 0.005, ** p-value > 0.01, *** p-value > 0.001

Supplemental Figure 5:

Upset plot showing the detection overlap between analyses in all cancer positive samples (A), late-stage cancers (B), early-stage cancers (C), breast cancers only (D), and cancer negative samples (E).

F) Comparison of genomic variants, fragment size z-score, and methylation signature across all patients with serial samples.

## Supplemental Tables

Supplemental Table 1 – Panel Design

## Notes

### Funding Statement

This study was funded by The Terry Fox Research Institute (grant number #1081) and Canadian Institutes for Health Research. This study was a collaboration with the CHARM consortium and is performed under the auspice of the LIBERATE study. Additional funding support was made possible by the Princess Margaret Cancer Foundation and the Ontario Institute for Cancer Research.

### Author Declarations

This study was approved by the University Health Network institutional review board (REB# 19-6239)

