## Supplementary figures and images for "Integrated analysis of cell-free DNA for the early detection of cancer in people with Li-Fraumeni Syndrome"

### Supplemental Figure 1

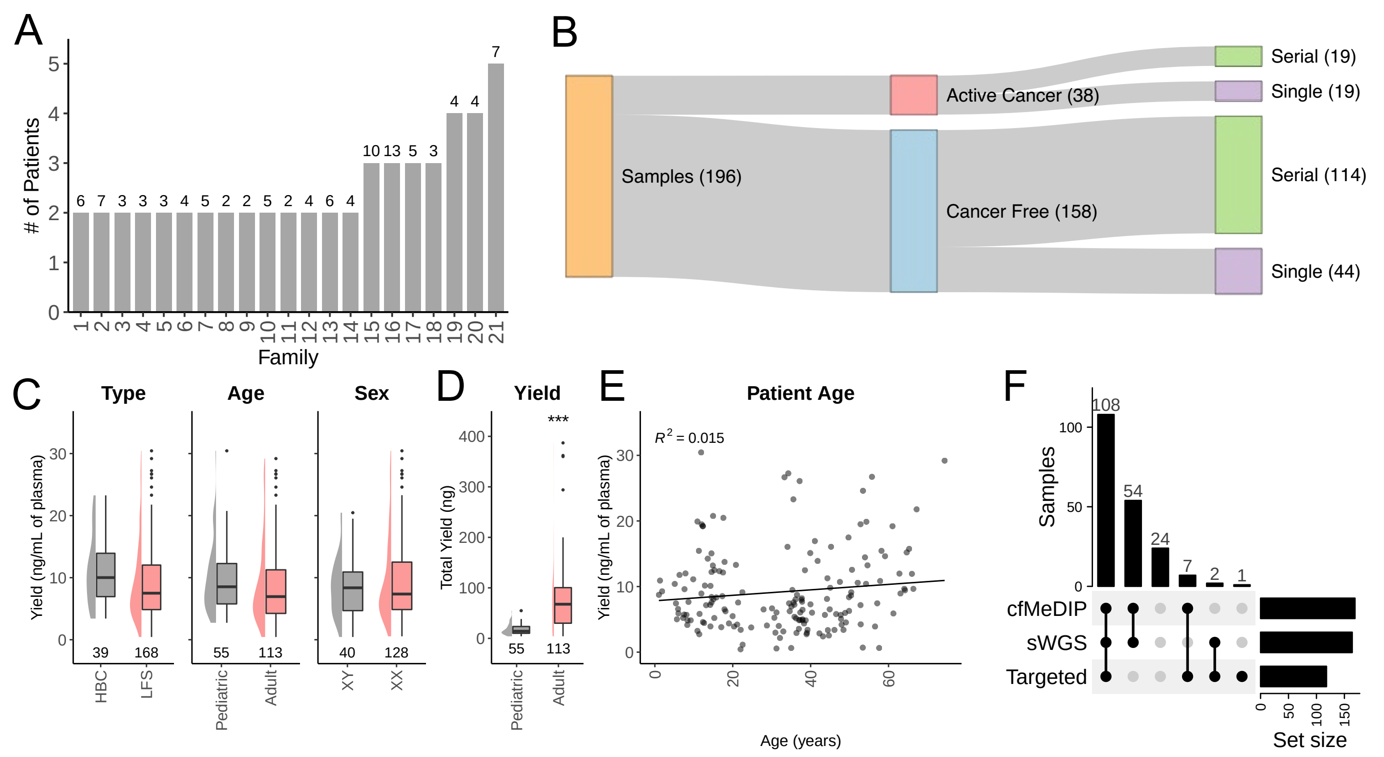

### Supplemental Figure 2

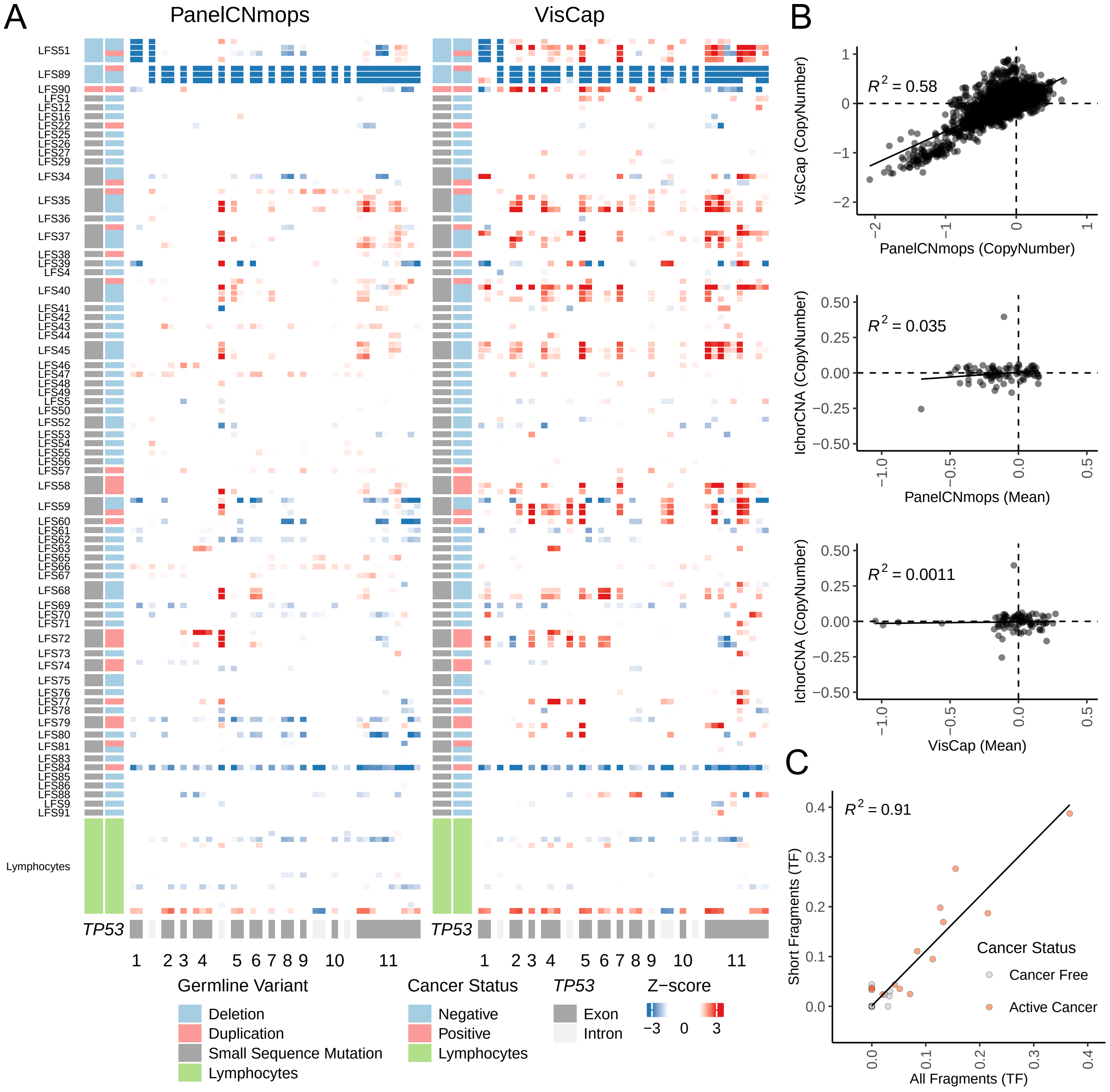

### Supplemental Figure 3

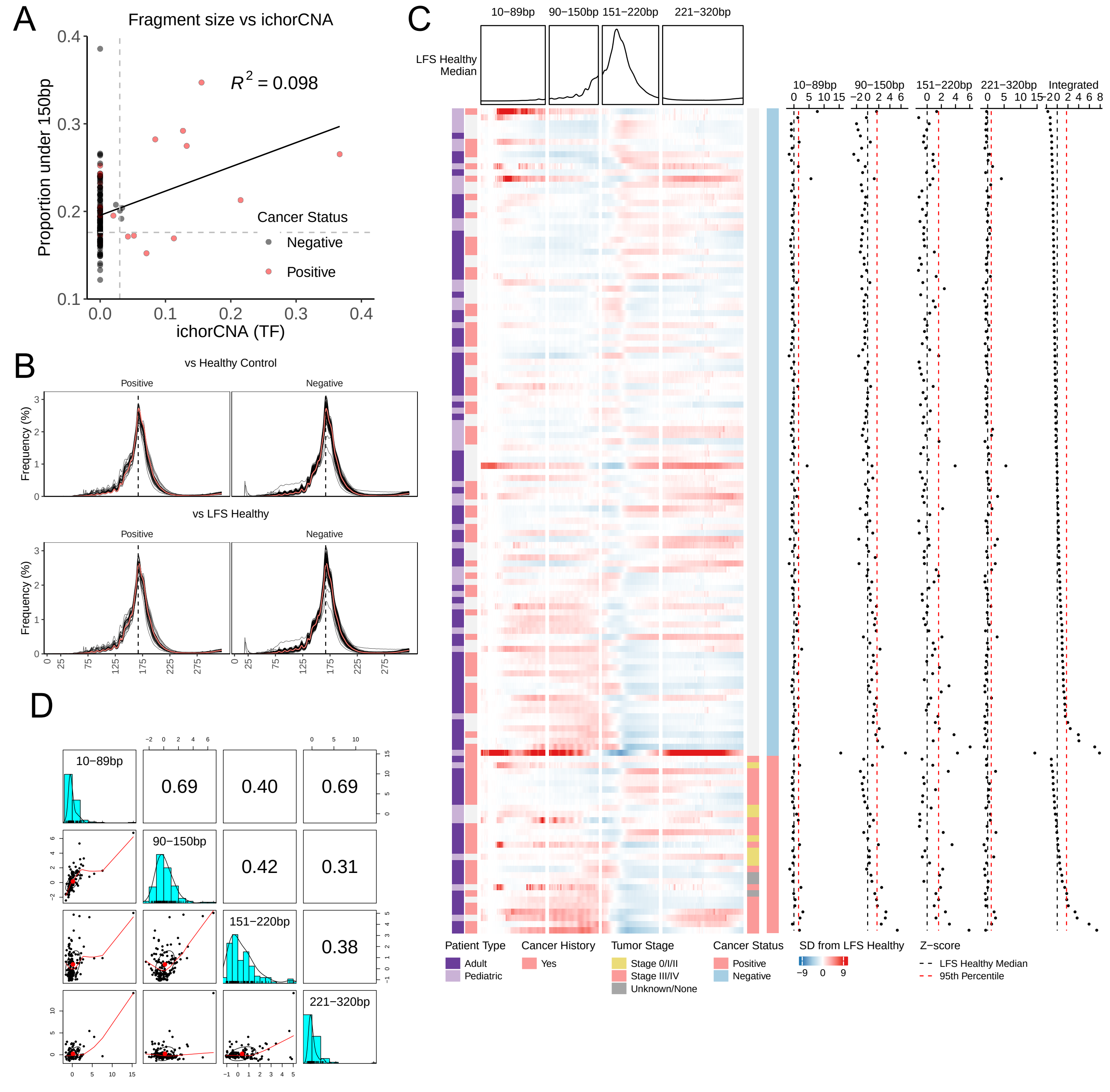

### Supplemental Figure 4

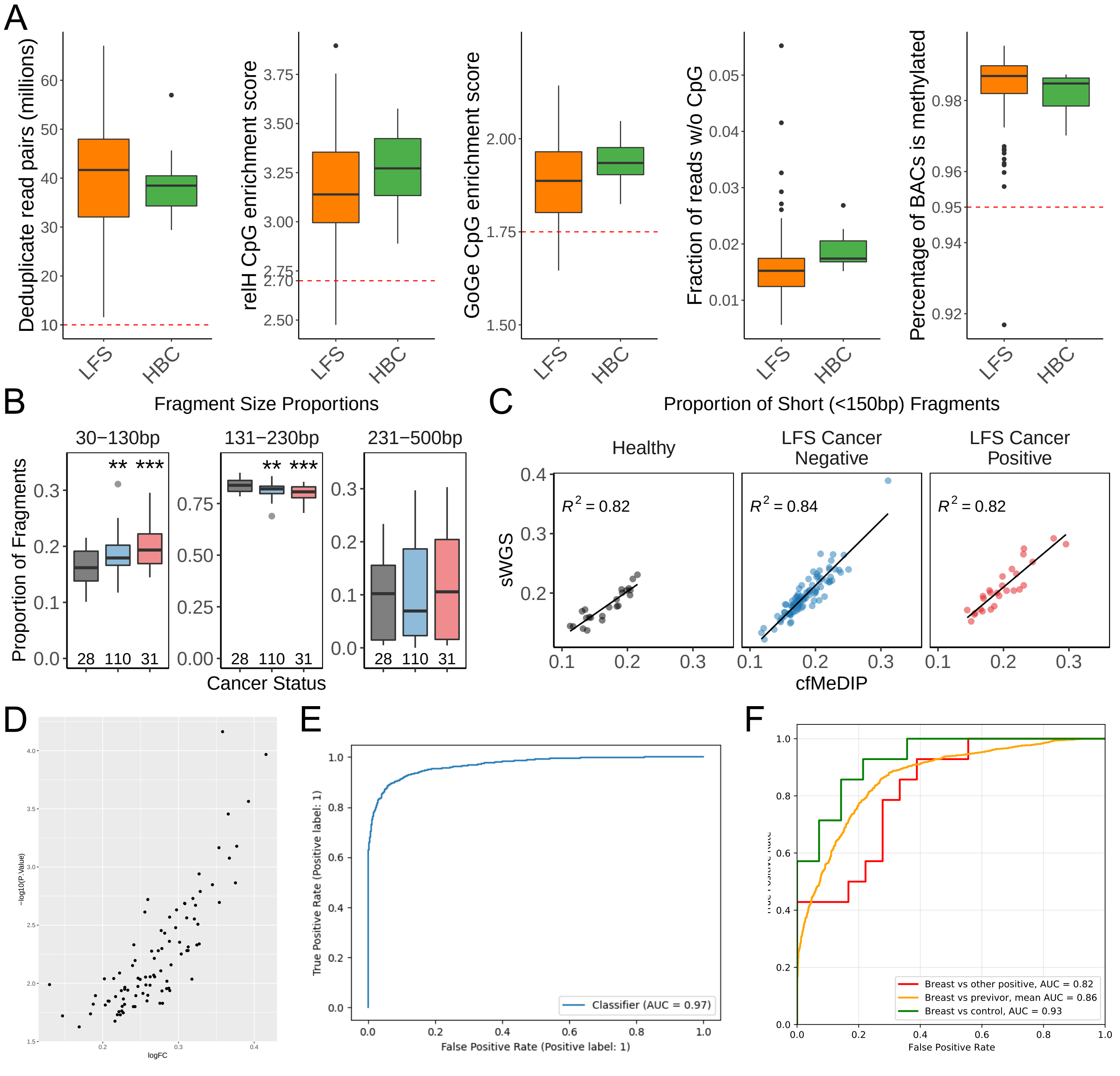

### Supplemental Figure 5

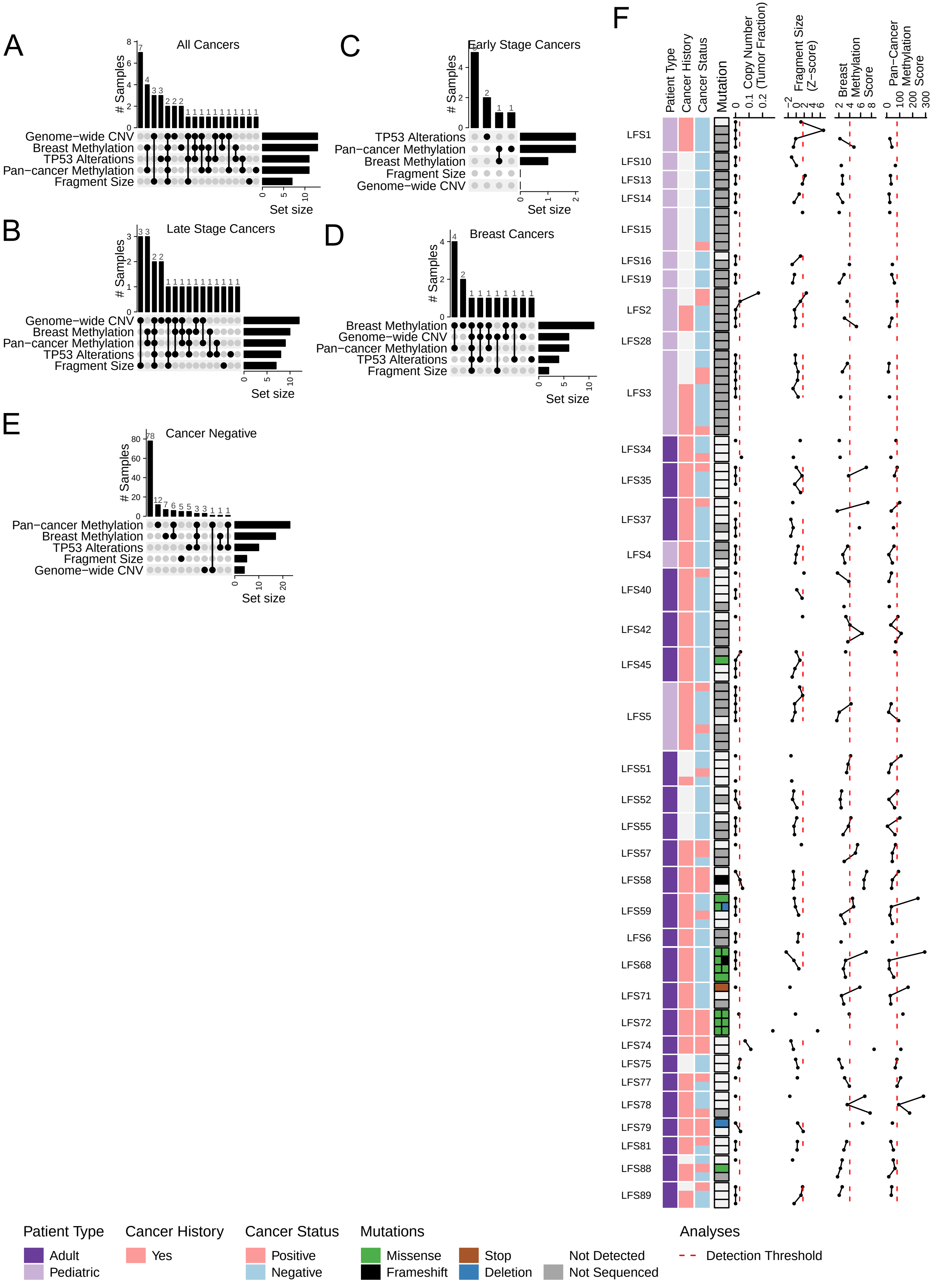
